# Factors influencing uptake of COVID-19 diagnostics in Sub-Saharan Africa: a rapid scoping review

**DOI:** 10.1101/2024.06.03.24308387

**Authors:** Mackwellings Maganizo Phiri, Yasmin Dunkley, Elizabeth Di Giacomo, Wezzie Lora, Moses Kumwenda, Itai Kabonga, Elvis Isere, John Bimba, Euphemia Sibanda, Augustine Choko, Karin Hatzold, Liz Corbett, Nicola Desmond

## Abstract

**Background:** Diagnostics are critical for preventing COVID-19 transmission, enabling disease management and engagement with care. However, COVID-19 testing uptake remained low in low- and middle- income countries in Sub-Saharan Africa (SSA) during the recent pandemic, due to issues of supply, access and acceptability. Early studies conducted outside of the region provide insight into uptake of COVID-19 testing, however there has been no systematic research within the region. The aim of this scoping review is to investigate factors influencing uptake of COVID-19 testing in different settings across SSA.

**Methods:** Inclusion criteria was any study employing qualitative or mixed methodologies, addressing uptake of COVID-19 testing conducted in SSA. MEDLINE, PubMed, Google Scholar, Web of Science, and Africa-Wide Information were searched.Thematic content analysis was conducted across all included articles until saturation was attained.

**Results:** In total 2994 articles were identified and fourteen reviewed. Structural, social, epidemiological, informational, and political elements affected how publics interacted with COVID-19 testing. Coverage was limited by insufficient diagnostic capabilities caused by a shortage of laboratory resources and trained personnel. False information spread through social media led to testing misperceptions and apprehension. Testing hesitancy was ascribed to fear of restrictive measures and the possibility of social harms if positive. Facility-based testing was physically inaccessible and perceived as lacking privacy, whereas self-testing distributed by the community removed lengthy distances and prevented stigma. Perceptions that COVID-19 was not severe and low numbers of confirmed cases in comparison to other settings undermined public urgency for testing. Low testing frequency led to low-rate assumptions, which in turn generated denial and othering narratives. Politicians’ acceptance or denial of COVID-19 affected the mobilization of the health system, and their model actions—such as testing openly—promoted public confidence and involvement in interventions.

**Conclusions:** This review emphasizes the necessity of strong political commitments to enhancing health systems for future pandemic preparedness. Response plans should consider contextual elements that affect how people react to interventions and perceive health emergencies. Community-driven self-testing distribution could enhance the uptake of diagnostics through addressing socio-economic constraints impacting facility-delivered testing.

## Introduction

Coronavirus disease (COVID-19) was declared a Public Health Emergency of International Concern by the World Health Organization (WHO) on January 30, 2020 [1,2]. Increased availability of diagnostic interventions for COVID-19 (C-19) was identified as a research priority, including delivering point-of-care (POC) testing within communities [2]. The WHO recommended integrating C-19 testing within routine diagnostics for other respiratory illnesses including influenza and tuberculosis to increase access [3]. Following these recommendations, different diagnostic techniques, including rapid diagnostic tests (RDTs), were produced and implemented [3]. These included genome sequencing, antigen or antibody detection, and molecular testing using nucleic acids [4]. Antigen/antibody tests were recommended for pandemic monitoring since they allowed rapid, regular, and expanded testing with onsite detection and immediate management [4]. Despite this potential, C-19 testing was not widely adopted by the public, particularly in low- and middle-income countries (LMIC) in sub-Saharan Africa (SSA) [5].

Identification of infected individuals through diagnostics is essential for disease prevention and control but testing-related challenges have been reported worldwide [5–8]. As C-19 spread, demand for diagnostic tests outstripped global supply, resulting in an inequitable access [5,6]. Although high-income countries had the means to produce or purchase technologies, access was limited in the LMIC [5,6]. This has been due to political and supply-side issues, including issues of global governance and health system-related factors such as resource limitations and logistics, as well as social and community-level factors such as communication and trust in delivery agents. The spread of misinformation undermined public confidence and restricted testing uptake globally [2]. Effective political leadership was demonstrated to impact engagement in preventive measures such as a sharp increase in people’s trust and willingness to test for C-19 when the president of Ghana tested publicly [9]. Likewise, where the political leadership was unwilling to test and dismissive of C-19 threat the desire to test among the general public was also correspondingly low.

Although studies have shed light on factors influencing public testing uptake, there has been little research in SSA specifically. User focus in SSA has been on general knowledge, attitudes, perceptions, and practices towards C-19 and vaccination responses, whilst supply-side research has investigated healthcare system conditions necessary for deploying testing instruments such as RDTs [10,11].

This study formed part of the “STAR Africa, Asia, Americas COVID-19 Preparedness Project (3ACP)” funded through UNITAID, investigating COVID-19 professional use and self-testing rapid diagnostics in Nigeria, Zimbabwe, and Malawi. As part of this work, we conducted a scoping of the contextual factors influencing people’s decisions regarding COVID-19 testing in various settings throughout SSA. This information would support the implementation of the main project.

## Methods

### Review scope

We conducted the review between July and August 2023. The review methodology is available at https://osf.io and has been registered with the Open Science Framework (OSF). Arksey and O’Malley’s methodological approach was used to formulate the research question, find relevant studies, choose studies, chart the data, and compile, summarize, and present findings [12]. Papers were selected following the Preferred Reporting Items for Systematic Reviews and Meta-Analyses (PRISMA) framework [13].

Inclusion criteria were any peer-reviewed study investigating factors influencing self- and provider-delivered COVID-19 testing uptake in sub-Saharan Africa with qualitative research methods described (i.e., focus groups, interviews, ethnography, and case studies), as well as mixed-methods studies including qualitative research conducted in conjunction with clinical trials. The types of diagnostic tests being used were another area of focus for data extraction. Quantitative research, literature reviews, and duplicates were removed from analysis after title screening. The search focused only on research from SSA from the onset of the C-19 pandemic (January 2020) to July 2023, when the review was conducted (**Table 1**).

**Table 1:**
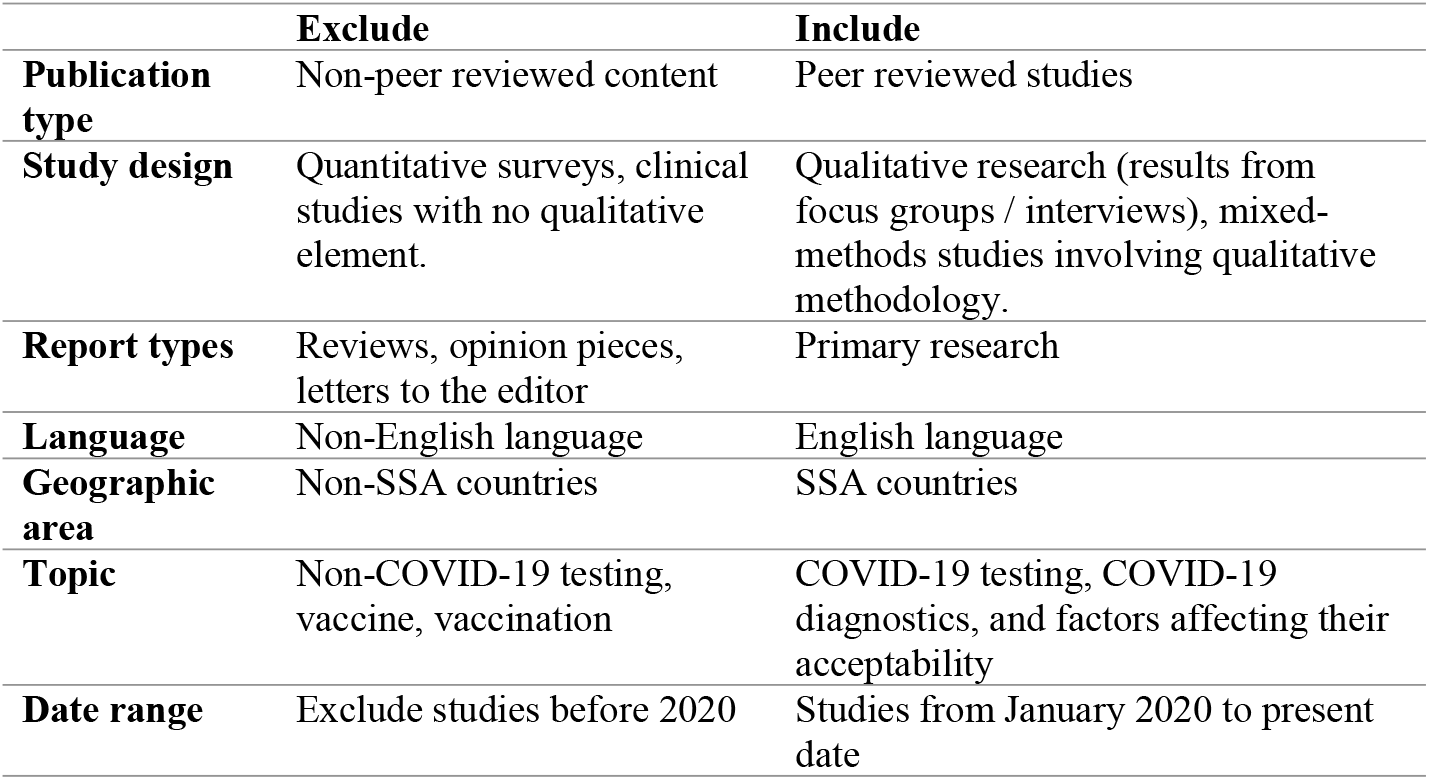
Inclusion and exclusion criteria

|  | **Exclude** | **Include** |
| --- | --- | --- |
| **Publication type** | Non-peer reviewed content | Peer reviewed studies |
| **Study design** | Quantitative surveys, clinical studies with no qualitative element. | Qualitative research (results from focus groups / interviews), mixed-methods studies involving qualitative methodology. |
| **Report types** | Reviews, opinion pieces, letters to the editor | Primary research |
| **Language** | Non-English language | English language |
| **Geographic area** | Non-SSA countries | SSA countries |
| **Topic** | Non-COVID-19 testing, vaccine, vaccination | COVID-19 testing, COVID-19 diagnostics, and factors affecting their acceptability |
| **Date range** | Exclude studies before 2020 | Studies from January 2020 to present date |

We conducted the search through Google Scholar, PubMed, Web of Science, Medline, and Africa-Wide Information. Search terms including COVID-19, COVID 19, coronavirus, testing, screening, RDTs, diagnostics, diagnose, enablers, facilitators, motivation, influence, behaviour, attitude, perception, beliefs, cultural, political, sociocultural, economic, social science, qualitative, and mixed methods were used. We used them separately as well as in combination (using the Boolean operators “AND” and “OR”) (Table 2). Filters were used to narrow the search to primary research abstracts and titles (**Table 2**).

**Table 2:**
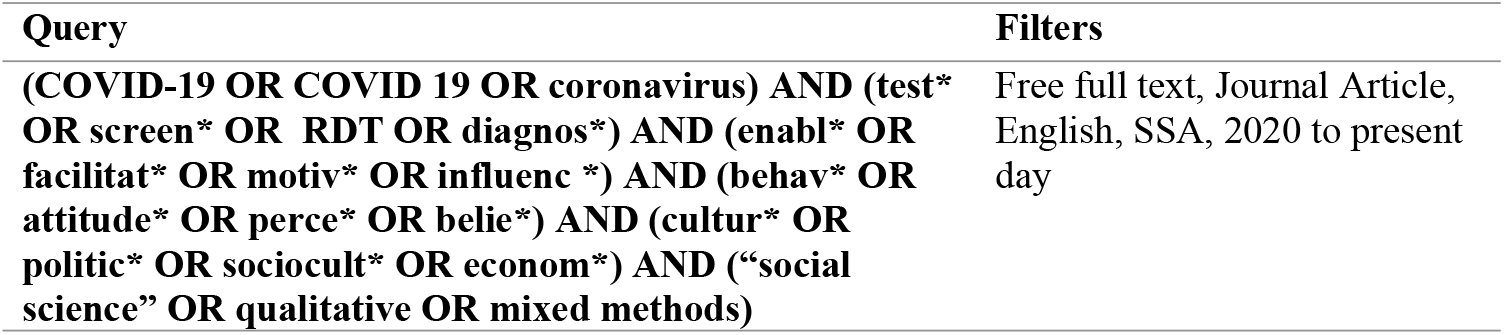
Search terms

Studies were initially included for consideration based on the title and abstract. If the abstract did not contain relevant information, we searched the article for the keywords described above. We evaluated the quality and relevance of eligible studies using a research appraisal tool developed by Hawker et al., 2002, and colleagues [14] (**S2**).We developed a data charting form including details on the author, publication year, location, study design, sample size, and conclusions **(S3)**.

## Data analysis

NVivo version 12 was used to import all the studies that satisfied the inclusion criteria. Codes and concepts were explored inductively and deductively. A preliminary coding framework was created and modified inductively to incorporate emerging themes. The initial codes and concepts were later reclassified, summarised, and integrated into two broad thematic areas: COVID-19 testing facilitators; and COVID-19 testing barriers. We present the data under these thematic categories, and **Fig 2** summarises the main and sub-themes under each category.

## Results

A total of 2994 studies were identified through the initial search across all databases. 2870 studies were eliminated. We screened the abstracts of 124 articles: 104 were excluded, covering topics related to COVID-19 but not directly associated with testing uptake, for example, COVID-19 vaccination, knowledge, and beliefs. Other reasons for exclusion included not focusing on the relevant disease area (HIV or tuberculosis diagnostics), while others were not conducted within SSA. We remained with 20 articles for full-text screening. Of these two were systematic reviews, three did not include qualitative approaches, and one was not conducted in SSA. A total of 14 articles remained for quality evaluation and data extraction (**Fig 1**).

**Fig 1.**
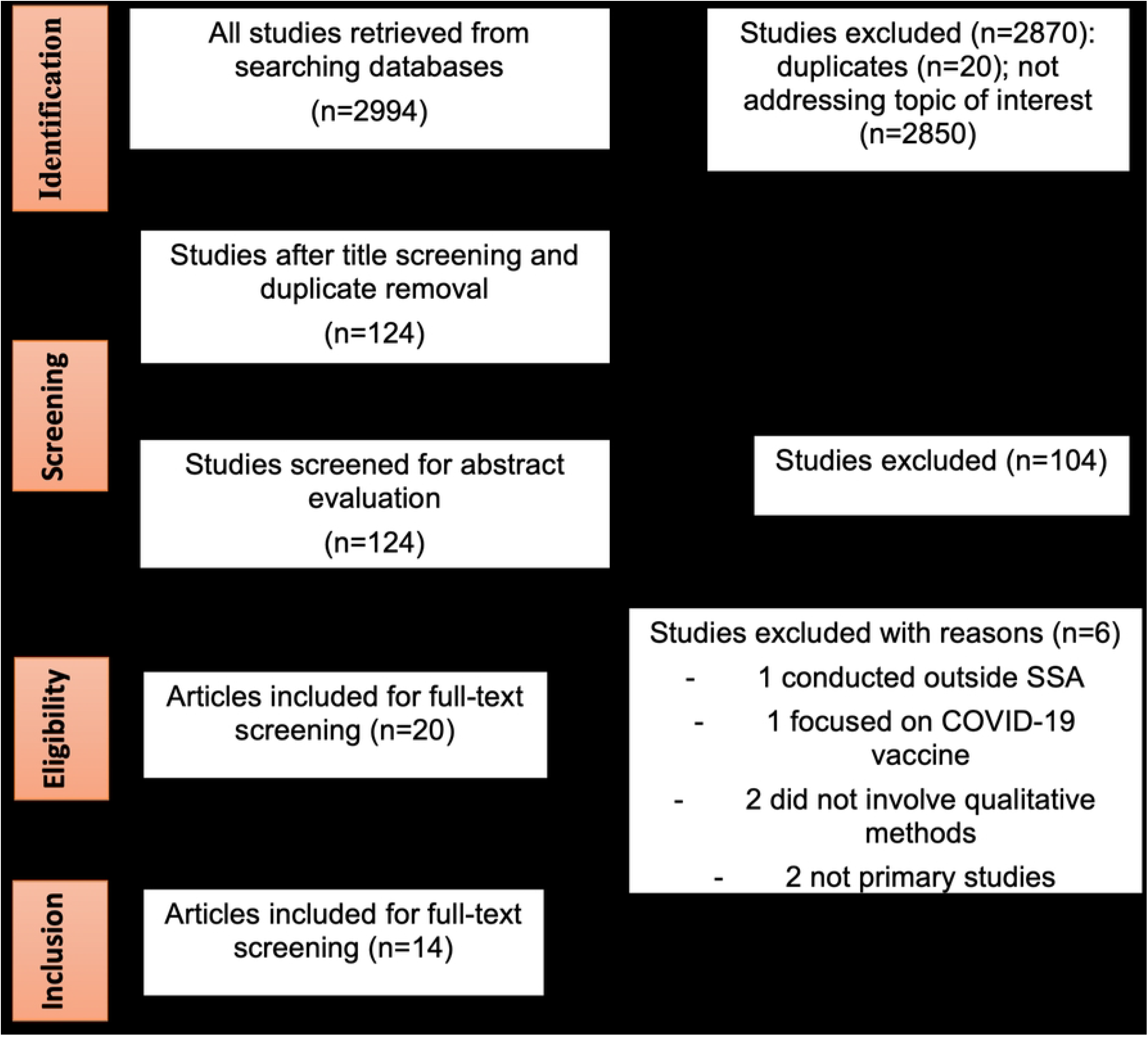
Process flow diagram for the research selection using PRISMA methodology.

**Fig 2:**
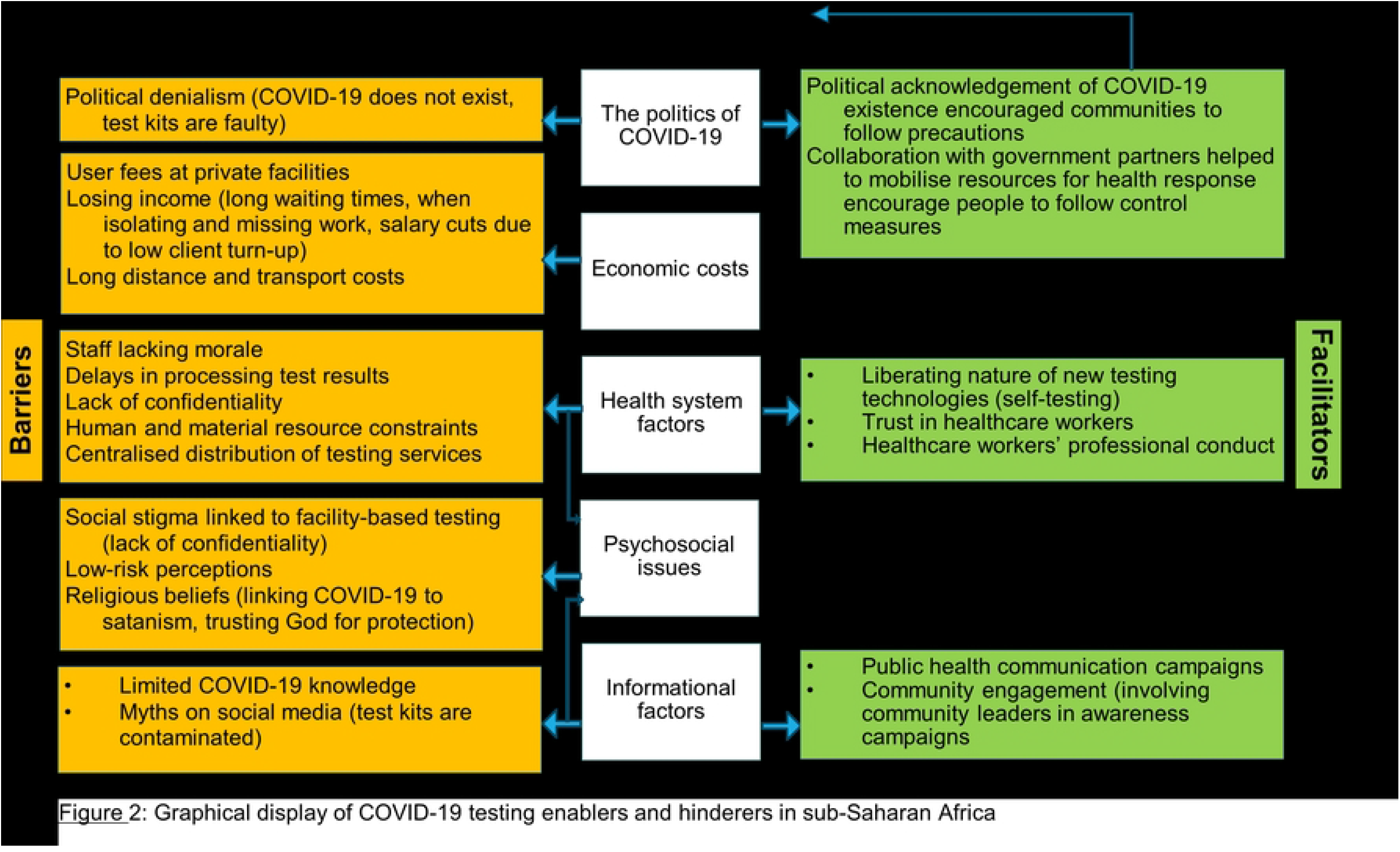
Graphical display of COVID-19 testing facilitators and barriers in sub-Saharan Africa.

Six studies analysed patient and stakeholder perceptions and experiences with C-19 testing and screening procedures [10, 16-20]. The remaining eight studies explored C-19 responses generally as well as testing-related topics. Four studies reported COVID-19 self-testing [9,15–17], five used facility-based RDTs [9,18–21], two used molecular tests [22,23], one used PCR tests [17], one used imaging [24], and two did not explicitly specify the diagnostic test used [25,26]. The studies had a total of 953 participants, aged 17 to 77 (**Table 3**).

**Table 3:**
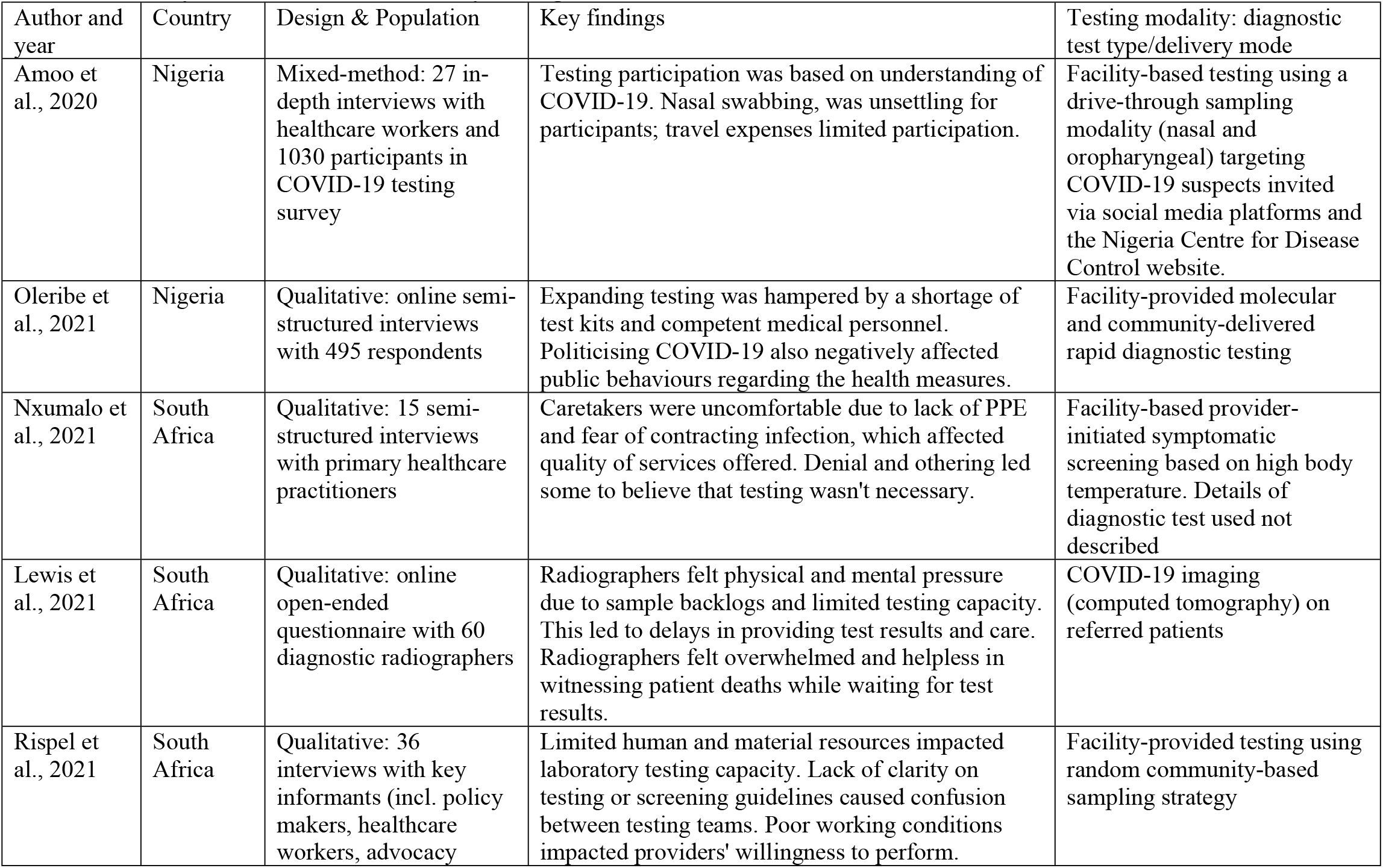
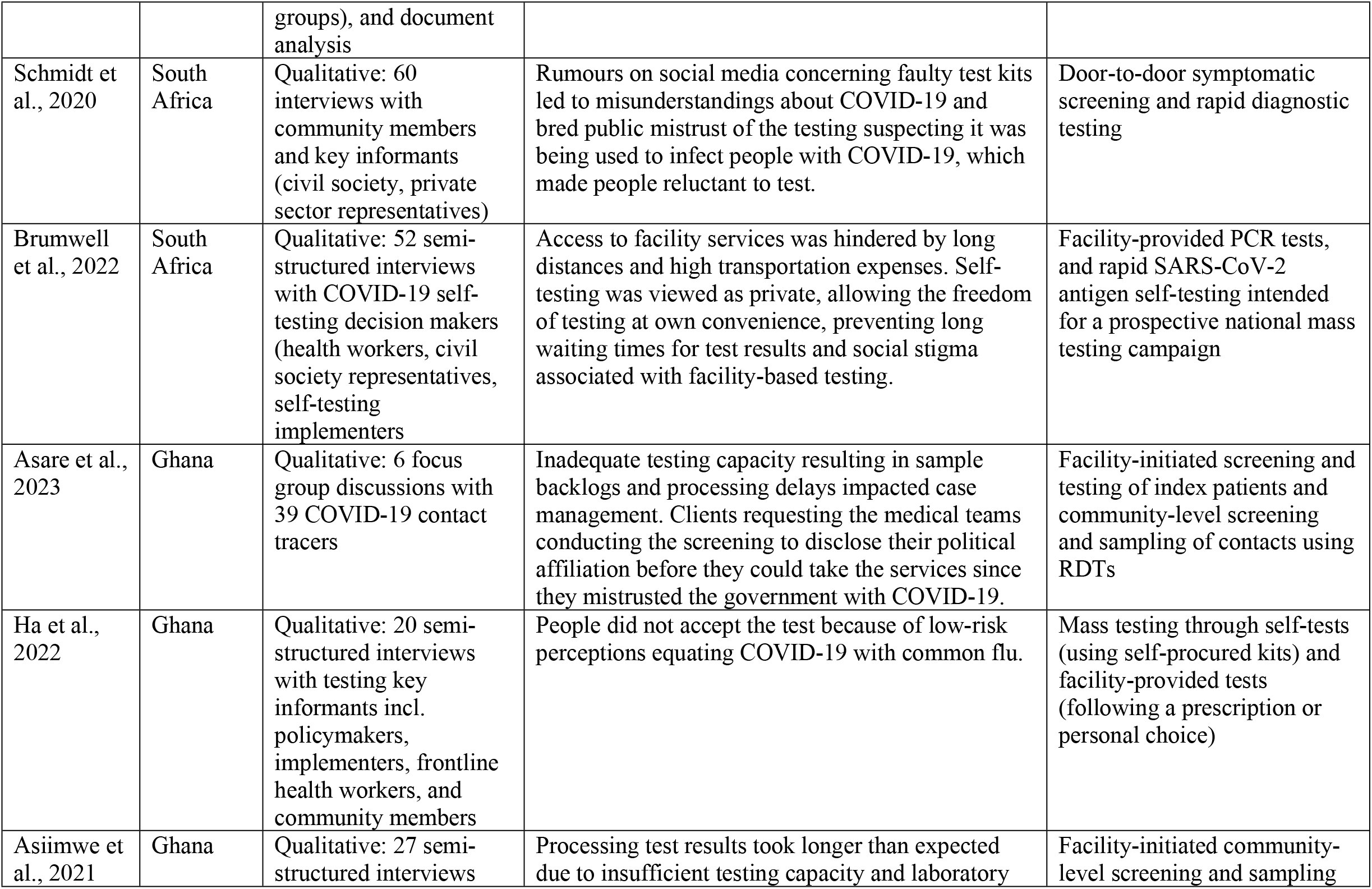
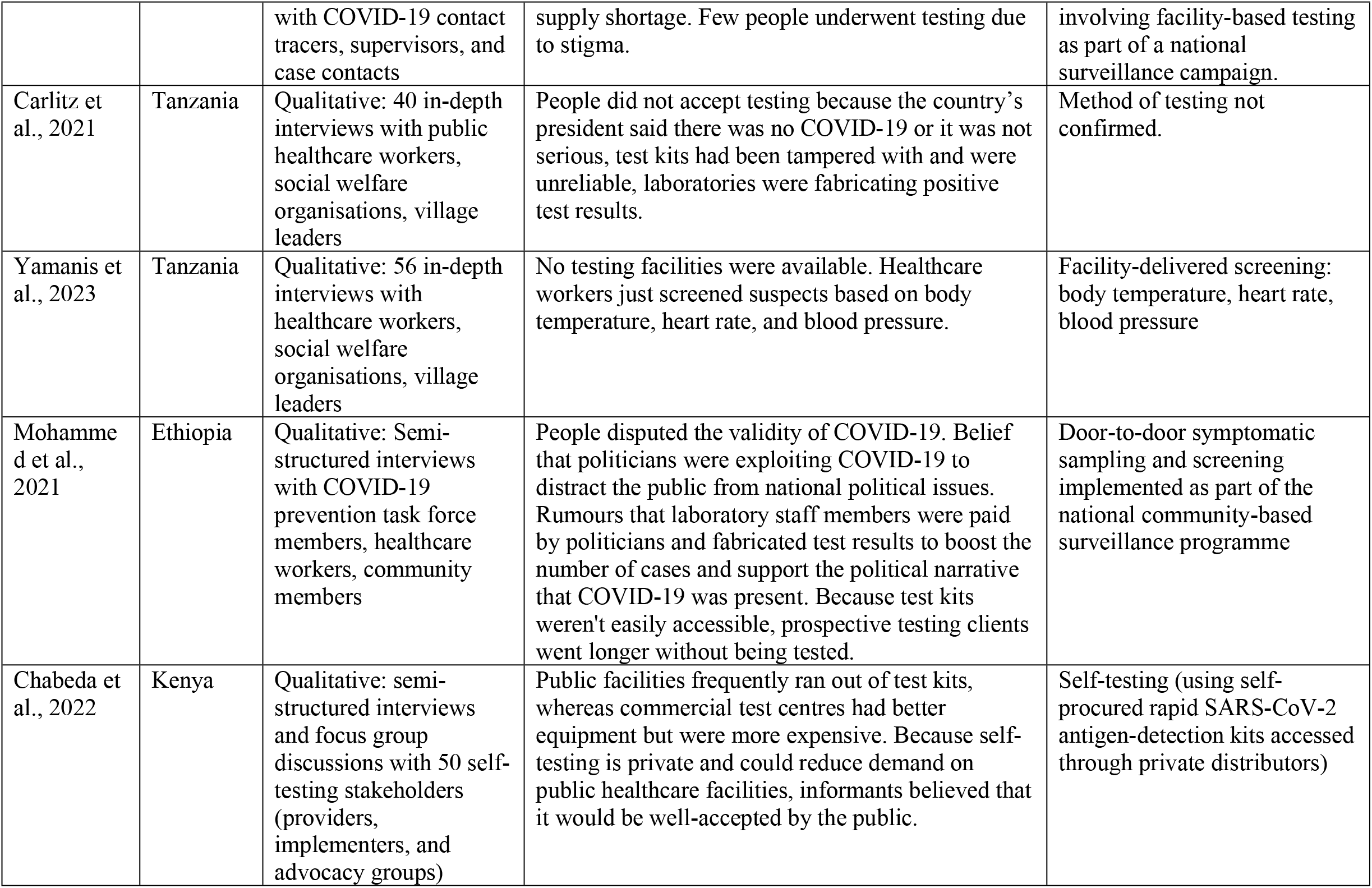
Summary of included studies with key findings

COVID-19 diagnostic uptake in SSA was problematic across many settings and influenced by political, institutional/structural, social, and informational factors.

### Facilitators of COVID-19 Testing

#### Effective political leadership

Strong political leadership was critical in determining the direction of the national COVID-19 response. Ha et al., 2022 and Yamanis et al., 20233 described that in areas where government officials viewed COVID-19 as a threat to public health, there was a strong political commitment to develop and implement disease containment measures including diagnostics. For example, in Ghana, the government gathered financial and material support to increase its diagnostic capacity through multisectoral partnerships with development partners. This allowed the country’s health system to expand the number of COVID-19 testing facilities nationwide, improving access and coverage of testing services, according to one of the nation’s laboratory managers:

> *“We did not have enough testing centres and PPE at the beginning of the pandemic. But, now, we have enough facilities, adequate PPE, and other consumables supported by the Ghana government, international organizations, and other donors for COVID-19 testing*.*”* [9]

Yamanis et al., 2023 described Tanzania making a similar commitment to empowering the health system and acknowledging the existence of COVID-19 after a period of denial when the new president, Hassan, acknowledged COVID-19 as a public health emergency relying on collaboration with local and international partners to improve control measures, including promoting testing uptake.

Both Ha et al., 2022 and Yamanis et al., 2023 described the influence of government in public responses to COVID-19 services. Political leaders not only made investments in health system capacity for COVID-19 monitoring activities, but took on a pro-public health advocacy role, urging people to get tested as well as follow the rest of the controls set in place. Some government representatives underwent COVID-19 testing or vaccination in public to legitimize and encourage improved public response, motivating testing:

> *We received lots of hope from the government and the president. Our president was really keen on tackling the pandemic […] We were highly encouraged to get tested by the president, and his leadership uplifted the motivation of getting tested*.*”* [9].

#### Public confidence in organizations providing testing

Public perceptions of the organizations tasked with carrying out testing activities were central to participation. In South Africa, Brumwell et al., found that people were more inclined to test for COVID-19 if they knew and trusted the providers:

> *“I think people do trust their pastors, their healthcare workers, nurses and general practitioners, pharmacists, principles…They generally don’t trust politicians. So, I wouldn’t include them there. But, generally, the community leaders, non-politically aligned, I think would be people that would be trustworthy*.*”* [17].

C-19 was more acceptable when spearheaded by people that community members were used to and had some form of prior association or interaction.

#### Novel COVID-19 testing modalities

Perspectives on self-testing were discussed in two studies by Brumwell et al., 2022 and Chabeda et al., 2022, and participants expressed a preference for self-testing over facility-based testing in both cases. Brumwell et al., 2022 claimed that this was due to testing flexibility, privacy, and confidentiality for socially excluded groups such as the homeless and drug users, whereas facility-based testing was felt to exacerbate stigma. Participants in Kenya in Chabeda et al., 2022 study described decision-makers perspectives that self-testing was crucial for physically vulnerable populations such as the elderly, the sick, those with impairments, and people living in isolated locations.However, they did not interview end users of self-testing.

. Knowledge of COVID-19 and its risks preceded the public adoption of testing. Eight of the studies reflected the usefulness of public health communication in generating demand for testing [9,16–18,20,23,25,27]. The studies frequently ascribed the uptake of testing to ongoing public health communication, which participants said was helpful in dispelling initial pandemic myths in the communities. Asare et al., 2023 mentioned that in Ghana finding contacts to test early in the pandemic was difficult, and awareness campaigns were seen to help communities respond more effectively.

### Barriers to COVID-19 Testing

#### Health system capacity

Nine of the studies identified that healthcare systems in SSA lacked the diagnostic tools and equipment necessary to identify COVID-19 patients, which impacted testing availability [9,17,18,20–23,25,26]. There was common recognition that systemic underfunding of healthcare systems, pre-existent to the pandemic, translated to a lack of preparedness for COVID-19. Problems included the paucity of test kits, disinfectants, and safety equipment in laboratories. Some hospitals lacked laboratory facilities altogether. For instance, in Tanzania Yamanis et al., 2023 revealed there was no laboratory to conduct testing, medical professionals merely checked suspects’ body temperatures, blood pressure, and heart rates. Inadequate laboratory readiness led to frequent backlogs in testing COVID-19 samples. Five of the studies reported protracted processing times ranging from several days [17,21,25,27] to more than a month [20] from sample collection, affecting COVID-19 public health response such as contact tracing.

#### Human Resource constraints

The availability of human capital also affected the availability of COVID-19 diagnostic testing in SSA. Four of the studies ([9,19,21,22] discussed the lack of skilled medical personnel to support testing and surveillance interventions. For example, a laboratory manager in Ghana described increased work burden due to staff shortages:

> *“We have only one person at the lab who runs the test. Despite our support, he ran samples until late. I also feel too exhausted […] when testing many people. The human personnel is fewer [*… ].*”* [9].

This shortage was also reported in South Africa and Nigeria [19,22]. Solutions suggested included retraining HIV service providers to reduce the supply-demand gap for COVID-19 testing.

Mental exhaustion and distress impacted staff capacity to respond to testing demands. Fears expressed included contracting COVID-19 through interaction with positive patients, where lack of access to personal protective equipment (PPE) and antiseptics for sanitizing surfaces was sub-optimal. Participants also felt underappreciated without support to address these issues. Fears were exacerbated when colleagues died of COVID-19. These factors all contributed to mental exhaustion.

#### Supply-Chain Issues

Five of the studies identified supply problems that impacted the availability and distribution of COVID-19 diagnostic services [9,17,20,21,23]. Mohammed et al. 2021 drew attention to budgetary constraints that impacted the purchase of medical necessities in Ethiopia, leading to inconsistent supply and frequent stockouts of test kits. Ha et al., 2022 described similar supply challenges such as irregular provision of personal protective equipment (PPE), making it difficult for surveillance teams to effectively conduct contact tracing:

> *“When COVID-19 [] came, we [the Ghana Health Service] were not prepared, which is why we faced a lot of challenges with contact tracing in the beginning. The PPEs were not there, yet we had to work. So, if the authorities could learn their lessons, I think we will be better prepared for the future*.*”* [9]

#### Accessibility of testing

Access to COVID-19 testing was geographically unevenly distributed across urban and rural settings. In one study, supply was better in urban centres than in rural ones, even when testing was supposedly available. Asare et al., 2023 in Ghana, for instance, described participants feeling that metropolitan facilities had more resources than their rural counterparts, making it simpler to receive services there. In this context, Brumwell et al., 2022 and Chabeda et al., 2022 demonstrated that self-testing could increase testing accessibility, helping to solve the issue of people failing to test because of large distances to facilities, which had an impact on both supply and demand. However, since patients had to travel to pick up the test kits, the supply was constrained by the central distribution of test kits through healthcare facilities.Participants in Brumwell et al., 2022 and Chabeda et al., 2022 believed that this posed the same challenges as facility-based testing.

#### Psycho-social and economic obstacles

Testing decisions were also shaped by risk perceptions and the economic and psychosocial ramifications of undergoing a test and being diagnosed with the disease. Seven studies [9,16–18,21,24,25] reported prevalent pandemic-related dread among community members, worrying about contracting and developing problems. Both Asiimwe et al., 2021 and Nxumalo et al., 2021 described these worries as stemming from social media rumours claiming that foreign locations had a high death rate. Carlitz et al., 2021 described that COVID-19 fatalities were being buried as Ebola victims, stories that increased fears of the pandemic and the social repercussions of receiving a COVID-19-positive diagnosis. In Brumwell et al., 2022 South African study, participants claimed that clients who tested positive experienced stigma because neighbours thought they were spreading the disease and held them responsible for new infections or fatalities.

Relating to economic costs, the two self-testing studies by Brumwell et al., 2022 and Chabeda et al., 2022 demonstrated that testing uptake was discouraged by the negative financial consequences of being diagnosed with COVID-19 and disclosure requirements. For the majority of those who tested positive, isolation requirements meant missing work. Failure to report for duty would also result in pay loss for jobs without sick days and participants believed that people’s fear of losing their income prevented them from testing and disclosing their status to prevent isolation. As mentioned earlier, transport costs incurred when accessing self-testing centrally distributed through facilities also dissuaded uptake [15,17].

#### False claims and beliefs

Nine studies reported how misinformation fuelled through social media encouraged negative perceptions of COVID-19, with a detrimental impact on demand for testing (Amoo et al., 2020; Asare et al., 2023; Brumwell et al., 2022; Carlitz et al., 2021; Ekohm et al., 2021; Ha et al., 2022; Mohammed et al., 2021; Nxumalo et al., 2021 and Schmidt et al., 2020). Mohammed et al., 2021 described a prevalent false claim that hospitals fabricated test results to increase the number of verified cases to demonstrate the reality of COVID-19.

Following the introduction of vaccination, rumours related to vaccines also impacted C-19 testing uptake. For example, Schmidt et al., 2020 highlighted refusal to uptake door-to-door screening and testing by medical personnel due to beliefs around vaccination in South Africa: *“Like as clinic staff we go in door-to-door, there are incidences where a house owner would refuse for us to go in, saying we don’t want your vaccines because they have Corona. Then we had to explain that we are not injecting people, we are just screening and asking questions. People are really scared, because of what they heard*…*”* [16]

Carlitz et al., 2021; Chabeda et al., 2022 and Schmidt et al., 2020 all described spiritual beliefs and religious beliefs that prevented the public from using tests and other interventions, compounding misconceptions spreading through social media. Chabeda et al., 2022 described belief in COVID-19 as a sign of devil worship in Kenya. Schmidt et al., 2020 described how COVID-19 was seen as testament that God was angry with humanity in South Africa.

#### Political exploitation of COVID-19 in SSA

Seven studies demonstrated how COVID-19 testing was highly politicized (Asare et al., 2023; Asiimwe et al., 2021; Carlitz et al., 2021; Mohammed et al., 2021; Oleribe et al., 2021; Yamanis et al., 2023). Studies in Tanzania by both Carlitz et al., 2021 and Yamanis et al., 2023 described political figures explicitly denouncing the pandemic’s existence, encouraging the public to seek herbal remedies. This, alongside the Tanzanian government’s decision to remove the country’s laboratory manager and end its monitoring program influenced willingness to adopt public health strategies including testing (Carlitz et al 2021).

The strength of perceived association between political agendas and C-19, fuelled through social media contributed to public mistrust of organizations providing health responses. In Ghana, Asare et al., 2023 highlighted the relationship between political affiliation and testing engagement where the public ‘screened’ providers of testing according to political views: “*Politicising the disease is a challenge to us [contact tracers]. This is because you will get to a contact’s home, and they start to politicise the entire process [of contact tracing] and they begin to ask you which party you belong to*.*”*

## Discussion

The findings of this review point to several structural, political, informational, economic, testing modality, and psychosocial elements that impacted directly on both provision and uptake of COVID-19 testing across SSA. Countries were unable to increase COVID-19 screening and testing because public healthcare systems lacked adequate laboratory and diagnostic equipment. The delivery of screening and testing was also influenced by safety worries and low morale among healthcare professionals because of a lack of protective equipment and compensation for additional work burdens. Demand and supply were both heavily impacted by political leadership. When effective this promoted resource mobilization, cultivated public trust, and encouraged participation in health interventions. In contrast, when government officials made COVID-19 a political issue, this bred mistrust and discouraged engagement. Willingness to test was influenced by perceptions of the professionalism of providers. Misinformation spread through social media related to vaccinations, politics, and testing outcomes, coupled with a lack of awareness about COVID-19 in general and the belief that this was a disease from elsewhere, were factors that tended to negatively influence views toward control measures.

Public testing choices were also affected by the nature of the test, the health dangers it posed, as well as its economic and psychosocial ramifications. For instance, people favoured self-testing over facility-based testing because the former required less travel time, offered testing liberty, ensured privacy, and lessened social stigma. The latter was unaffordable due to the great distance, expensive cost, and risk of disease transmission from traffic. Healthcare workers also preferred the self-testing modality because it helped to relieve health system burdens. However, COVID-19 self-testing was not key in most of the studies as only two examined perspectives on its acceptability.

Our findings are consistent with previous research, particularly relating to the factors that promote or impede the implementation and uptake of point-of-care diagnostic interventions for pandemics in SSA, including for HIV and Ebola. For instance, several studies have demonstrated limited public engagement with facility-based HIV testing because people felt the model involved long travels and was inconvenient, lacked privacy, caused stigma and discrimination, and limited their autonomy [28,29]. Relating to supply chain issues, a systematic study on HIV diagnostics in low-and middle-income settings including SSA identified the lack of laboratory equipment as one of the key factors undermining HIV testing programmes [30]. Similarly, shortages of medical equipment and resources hampered public health efforts during the 2014 Ebola outbreak in West Africa to identify those who were infected with the virus [31,32]. Agreeing with our results, a review of HIV testing enablers and barriers in Africa showed that self-care options such as HIV self-testing granted users the freedom and convenience of testing at the place and time of choice, reduced the stigma and discrimination associated with facility-based testing, and boosted HIV testing uptake [30].

The laboratory and diagnostic challenges highlighted by this research have significant effects on country-level ability to control infectious disease outbreaks. Epidemiological surveillance is also challenged when affected individuals go undetected, raising the risk of transmission, and making it more difficult to implement interventions in response to epidemics [33]. Governments may become more self-sufficient and better equipped for upcoming pandemics if domestic resource revenue is maximized under strong political leadership [31].

## Strengths and limitations

The study was enhanced by the systematic searching of several databases to find all relevant studies that satisfied the predefined inclusion criteria. Understanding of the variables influencing COVID-19 testing uptake was enriched through the inclusion of papers employing a variety of methodological techniques, including mixed-methods studies. Regarding limitations, restricting the inclusion of studies only to those published in English due to language barriers entailed a possibility of missing other relevant studies. The reviewed papers were written at specific time points, raising the possibility of the findings not reflecting the rapid changes in pandemic responses and how people reacted to them overtime. Primary studies addressing the research question were also scarce at the time of the review, and the few that we analysed examined COVID-19 testing largely from the viewpoints of decision-makers as opposed to actual testers. This remains a knowledge gap regarding the actual testing experiences, which would have deepened the analysis of the demand-side facilitators and barriers. To better understand uptake drivers and match testing outcomes with social contextual needs, future pandemic diagnostic testing research should prioritize end users.

## Conclusion

The COVID-19 pandemic response in SSA was dynamic and testing provision and uptake changed over time. Initially, many SSA countries lacked the resources to identify all COVID-19 cases [32] and it may be likely that cases were consequently underreported [33]. Healthcare systems had received little funding and lacked the equipment and personnel needed to efficiently prepare for and conduct testing. This emphasizes the necessity of a strong political commitment to enhancing health systems for pandemic preparedness in the future. Future pandemic response plans should consider contextual elements that affect how people react to interventions and perceive health emergencies. Self-testing solutions that are distributed by the community could remove socioeconomic constraints frequently associated with facility-delivered testing and increase access to pandemic diagnostic services. To ensure proper lay use of these self-care devices and linkage to care, user-friendly instructions and community-based psychosocial support networks are crucial factors.

## Data Availability

Data underlying this research were peer reviewed articles accessed through the internet. We have included in the submission notes from NVIVO coding together with to the articles that were included and analysed for the scoping review.

## Ethics

This investigation did not seek specific ethics approval because it analysed secondary data without involving primary data collection with human subjects. However, all the country-specific projects that it was part of received individual ethical approvals from in-country, the London School of Tropical Hygiene and Medicine, the Liverpool School of Tropical Medicine, and the WHO (S1).

## Acknowledgments

The authors would like to thank the entire 3ACP research group for supporting this work.

## Supporting Information

**S1**. Ethical approval numbers for in country 3ACP studies, informed through this scoping review.

**S2**. Completed study quality and relevance form.

**S3**: Completed data charting form

